# The broad impact of cell death genes on the human disease phenome

**DOI:** 10.1101/2023.06.11.23291256

**Authors:** Abigail Rich, Phillip Lin, Eric Gamazon, Sandra Zinkel

**Affiliations:** Molecular Pathology & Immunology Graduate Program, Vanderbilt University; Department of Medicine, Vanderbilt University Medical Center

## Abstract

Apoptotic, necroptotic, and pyroptotic cell death pathways are attractive and druggable targets for many human diseases, however the tissue specificity of these pathways and the relationship between these pathways and human disease is poorly characterized. Understanding the impact of modulating cell death gene expression on the human phenome could inform clinical investigation of cell death pathway-modulating therapeutics in human disorders by identifying novel trait associations and by detecting tissue-specific side effect profiles. We analyzed the expression profiles of an array of 44 cell death genes across somatic tissues in GTEx v8 and investigated the relationship between tissue-specific genetically determined expression of 44 cell death genes and the human phenome using summary statistics-based transcriptome wide association studies (TWAS) on human traits in the UK Biobank V3 (n ∼500,000). We evaluated 513 traits encompassing ICD-10 defined diagnoses and hematologic traits (blood count labs). Our analysis revealed hundreds of significant (FDR<0.05) associations between cell death gene expression and diverse human phenotypes, which were independently validated in another large-scale biobank. Cell death genes were highly enriched for significant associations with blood traits versus non-cell-death genes, with apoptosis-associated genes enriched for leukocyte and platelet traits and necroptosis gene associations enriched for erythroid traits (e.g., Reticulocyte count, FDR=0.004). This suggests that immunogenic cell death pathways play an important role in regulating erythropoiesis and reinforces the paradigm that apoptosis pathway genes are critical for white blood cell and platelet development. Of functionally analogous genes, for instance pro-survival BCL2 family members, trait/direction-of-effect relationships were heterogeneous across blood traits. Overall, these results suggest that even functionally similar and/or orthologous cell death genes play *distinct* roles in their contribution to human phenotypes, and that cell death genes influence a diverse array of human traits.

## INTRODUCTION

Regulated cell death is an essential phenomenon during the development of multicellular organisms^1–3^ and an inevitable consequence of cellular and organismal aging^2, 4^. Three well characterized forms of cell death that rely upon genetically encoded, hierarchical signaling pathways are apoptosis, necroptosis, and pyroptosis^5^.

Apoptosis can be elicited via extrinsic or intrinsic cellular perturbation; as a result, there are both “extrinsic” apoptosis regulated by caspases and “intrinsic” apoptosis regulated by BCL-2 family members which regulate mitochondrial membrane permeability^6, 7^. Necroptosis is a regulated form of cellular necrosis which converges on the assembly of an MLKL pore on the cell membrane^8, 9^. Pyroptosis is an immunogenic form of cell death that is canonically reliant upon the formation of the NLRP3 inflammasome and the release of IL-1beta and IL-18 via gasdermin membrane pores^10^. Apoptosis, necroptosis, and pyroptosis all ultimately result in cellular demise; however, their mechanisms and functions are unique.

### Three major regulated cell death modalities are implicated in health and disease

Regulated cell death is a homeostatic biological process and dysregulation of cell death has pathophysiological implications^11–13^. For instance, neurodegenerative disease is associated with inappropriate neuronal cell death and inflammatory cell death pathways^14^. Bone marrow failure syndromes are associated with the upregulation of necroptotic and pyroptotic inflammatory cell death programs within the marrow^15–17^. Neoplasms are associated with both the upregulation of prosurvival pathways and the downregulation of pro-death signals^18–20^; mouse models with both systemic and tissue-specific deficiencies in key apoptotic signaling regulators have illustrated a propensity for the development of malignancy^21–23^. Autoimmune diseases arise from defects in the control of autoreactive lymphocytes: TNFR-dependent extrinsic apoptosis is crucial for the negative selection of autoreactive thymocytes, and antiapoptotic BCL2 family members play a role in regulation of this process^24–27^; BCL2 family signaling contributes to selection against polyreactive B cells^28–31^. The Mendelian disease autoimmune lymphoproliferative syndrome (ALPS) arises from defects in the genes *FAS*, *CASP8*, and *CASP10*, which coordinate the Fas-dependent apoptosis that is critical for thymocyte development^32, 33^. As suggested by the above examples, defects in cell death regulation contribute to a diverse array of pathologies that can be tissue- and context-specific.

### Tissue-specific transcriptional regulation of cell death has implications for human disease

Diverse functions of various somatic tissues dictate distinct cell death behavior^34, 35^ despite developing from identical germline DNA. For example, skin cells must be resistant to cell death to perform their barrier function, whereas the dynamic regulation required of the hematopoietic system dictates a similarly dynamic regulation of cell death to maintain homeostasis. This context dependency is established in part by transcriptional regulation of cell death pathway members, which operate downstream of transcription factor programs including Rel/NF-kappaB^36^, p53^37^, interferon regulatory factors, among others^10, 38^. Gene expression patterning is generally tissue-specific and varies across individuals in a population^39, 40^. Although the role of cell death pathway genes has been rigorously defined during development in mouse models, the specific landscape of the expression of cell death pathway genes across human somatic tissues is a fundamental aspect of cell death biology and has not been rigorously characterized. In this study, we examine the relationship between tissue-specific expression of cell death genes to characterize the landscape of cell death gene expression across human tissues. From there, we examine the relationship between tissue-specific cell death gene expression and human traits by leveraging summary statistics-based transcriptome wide association studies (S-TWAS)^41^ to identify how subtle, lifetime shifts in the predisposition of cell death gene expression associates with clinically relevant traits across the human phenome.

## RESULTS

### Curation of an array of core cell death genes

Our studies focus on core programmed cell death machinery in the apoptotic, necroptotic, and pyroptotic pathways as they are well studied^5, 42^, targetable by multiple clinically relevant pharmaceuticals^43–45^, and therefore implicated in the pathogenesis of many diseases^11, 12, 46^. In all, we chose 44 genes operating within the intrinsic apoptosis pathway (18 genes), extrinsic apoptosis pathway (12 genes), necroptosis pathway (4 genes), and pyroptosis pathway (10 genes) (Fig. 1, Supp. Table 1).

**FIGURE 1:**
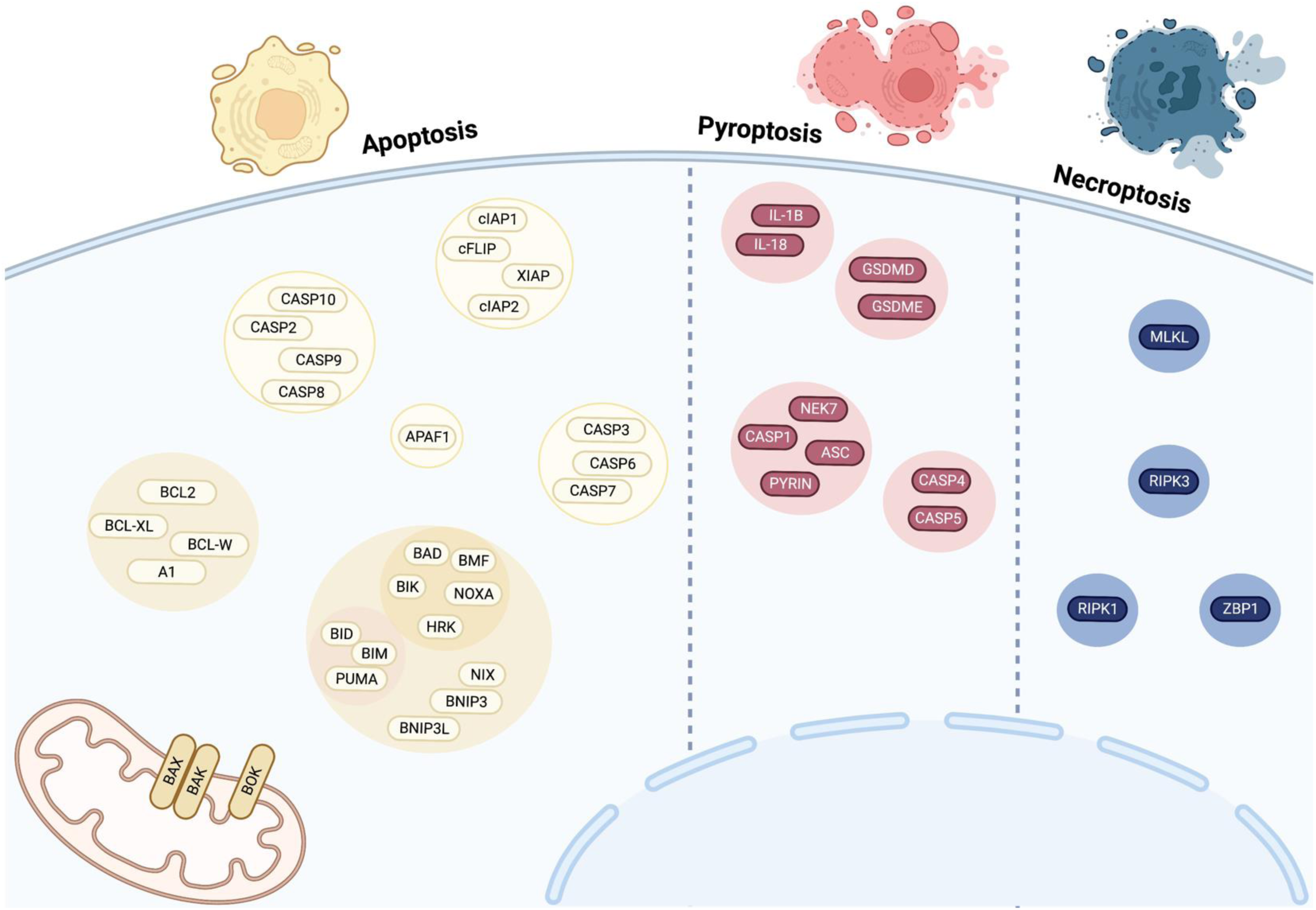
Composition and organization of an array of cell death genes. Summary of the composition and organization of our chosen cell death gene array by pathway and subpathway: three cell death modalities of interest: apoptosis (yellow), pyroptosis (red), and necroptosis (blue) and the proteins encoded by the genes selected for studying these modalities (ovals). Proteins are grouped by function.

### Observed expression of programmed cell death genes is variable and tissue-specific

We first examined the expression of members of our cell death gene array across adult somatic tissues to reveal cell-type-specific patterns in cell death machinery networks (Fig. 2A). We employed the GTEx resource release 8, which has transcriptomic data on somatic tissues for 838 individuals. Across 49 GTEx tissues, expression is highest in genes encoding prosurvival factors *MCL1* (MCL1), *BCL2L1* (BCL-XL), and *BCL2L2* (BCL-W) (Fig. 2B). There was variable expression of all these genes across tissues, which can be visualized by the interquartile spread when median log2(TPM) expression for each of these genes was graphed (Fig. 2B). The variance observed was not explained by the number of expression observations in GTEx v8, suggesting that these expression distributions are biologically relevant (rather than a sample size artifact) (Supp. Fig 1A). Two possible interpretations of this variance in cross-tissue gene expression are: expression patterns of cell death genes maintain a similar stoichiometry but differing magnitude across tissues; or expression patterns of cell death genes are variable across tissues.

**FIGURE 2:**
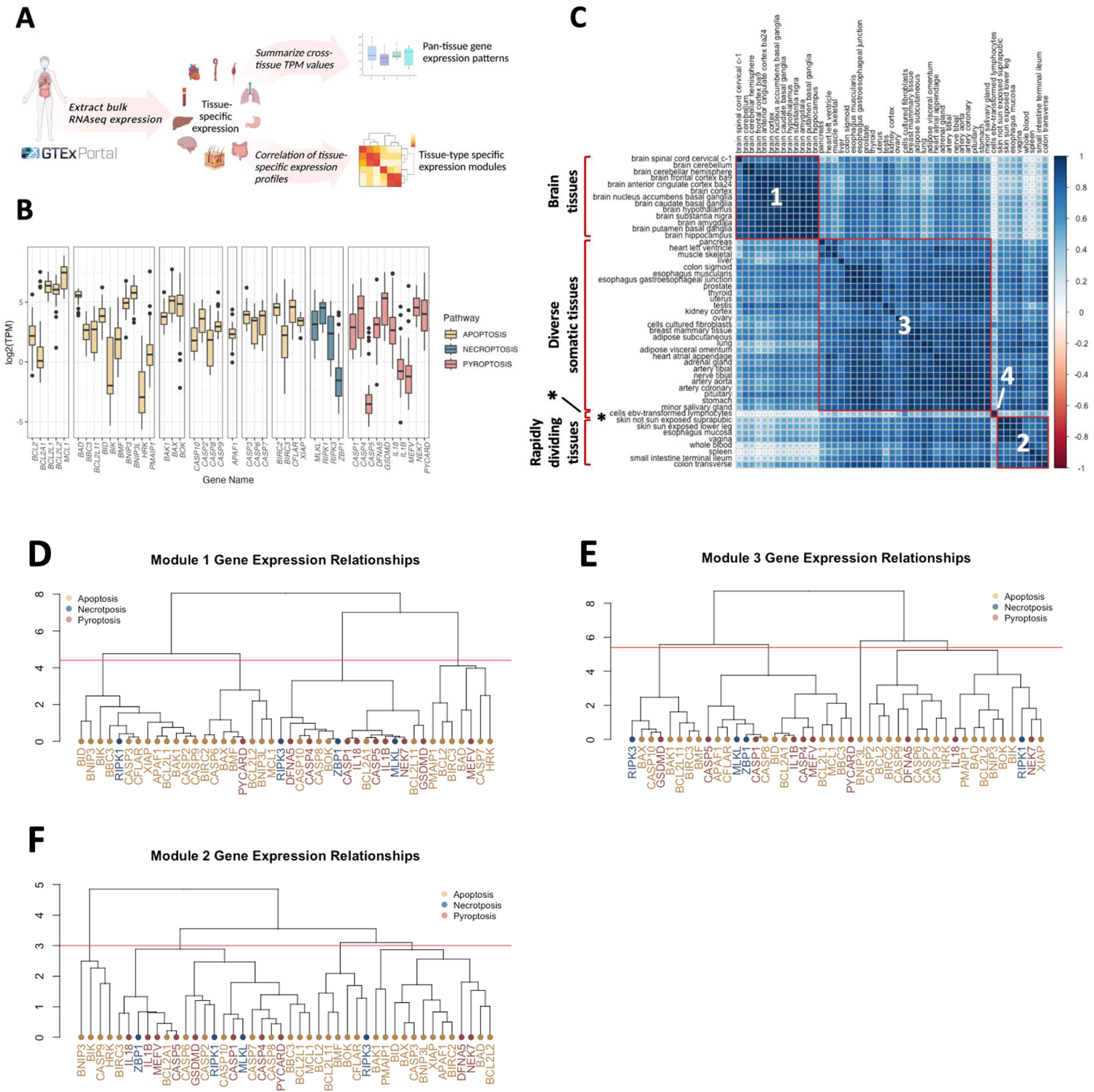
Tissue-specific patterning of cell death gene expression across adult somatic tissues. 2A: Preprocessed GTEx tissue expression data in transcripts per million (TPM) from 49 tissues were extracted for analysis, and the distribution of median values in each tissue and the Pearson correlation between median TPM values in each tissue was calculated. 2B: Boxplot depicting the distribution of median tissue TPM for GTEx tissues for each cell death gene. Outlier expression values (>1.5X IQR) are depicted as dots. Genes are grouped by pathway. 2C: Correlation plot for 49 GTEx tissues using Ward’s D hierarchical clustering reveals four modules (outlined in red) that are highly correlated: brain tissues, rapidly dividing tissues, diverse somatic tissues, and EBV-transformed lymphocytes (indicated by “*”). 2D-F: Dendrograms illustrating relationships between gene expression values in Modules 1 (D), 2 (E), and 3 (F) using hierarchical clustering of Euclidean distance between median gene TPM values across members of expression modules identified in (C).

To ascertain if gene expression patterns were maintained across tissues, we computed the correlation of each tissue-tissue pair using expression of all genes in our cell death array. Hierarchical clustering of correlation values subsequently identified four distinct groups of tissues with highly correlated expression of cell death genes (Fig. 2C). Interestingly, these tissues segregated into biological themes as follows: brain tissues, rapidly dividing tissues, diverse somatic tissues, and transformed lymphocytes (Fig. 2C, Supp. Table 2). These tissue modules have distinct gene co-expression patterns. The correlation coefficients derived from examining the relationship between median expression of each cell death gene-gene pair across all tissues in a module ranged widely (from ρ = -.99 to 0.99) (Supp. Fig. 1A, Supp. Table 3). These module-specific expression patterns suggest differences in “wiring” of cell death pathways across cell and tissue types. Module-specific gene expression patterning reveals that the most highly related genes transcend classical pathway boundaries (Fig. 2D-F). The distinct patterns we observed highlight the importance of evaluating expression in a tissue- or module-specific context for understanding tissue and organ-specific cell death dynamics. Furthermore, the diverse tissue-specific expression patterns we observed suggest that specific human diseases might be associated with tissue-specific changes in cell death gene expression.

### Transcriptome-wide association studies identify associations between genetically determined expression of cell death genes and human disease

#### Joint Tissue Imputation (JTI) generates prediction models for most cell death genes across human tissues for large-scale use on GWAS summary statistics

To address the relationship between tissue-specific genetically determined expression of cell death genes and human disease on a tissue-specific level, we implemented a unique summary statistics transcriptome-wide association study (TWAS) approach, Joint Tissue Integration (JTI) methodology^47^, on 49 separate GTEx v8 tissues (Fig. 3A, Supplemental Table 4). Joint tissue imputation generated *in silico* genetic variation based models of gene expression for 43 of our array of 44 cell death genes involved in apoptosis, necroptosis, and pyroptosis. Two genes, *CASP7* and *BAK1*, had sufficient QTL information or gene expression heritability for testing in all 49 GTEx tissues, whereas *XIAP* and *BCL2A1* were included in zero and two tissue-models, respectively (Supp. Table 5, Supp. Fig. 2A). This observation suggests that cell death genes may have variable level of genetic control of expression across tissues. These models can then be applied to GWAS summary statistics to estimate the association between genetically-determined gene expression and human traits. Because the level of genetic control varies with tissue, certain genes are tested more frequently for each trait than others in these analyses, and some gene/tissue relationships are not tested. At least 10 genes were tested in each tissue (Supp. Fig. 2B). Overall, these models test for associations between 1,061 cell death gene/tissue pairs and any trait of interest for which GWAS summary statistics are available.

**FIGURE 3:**
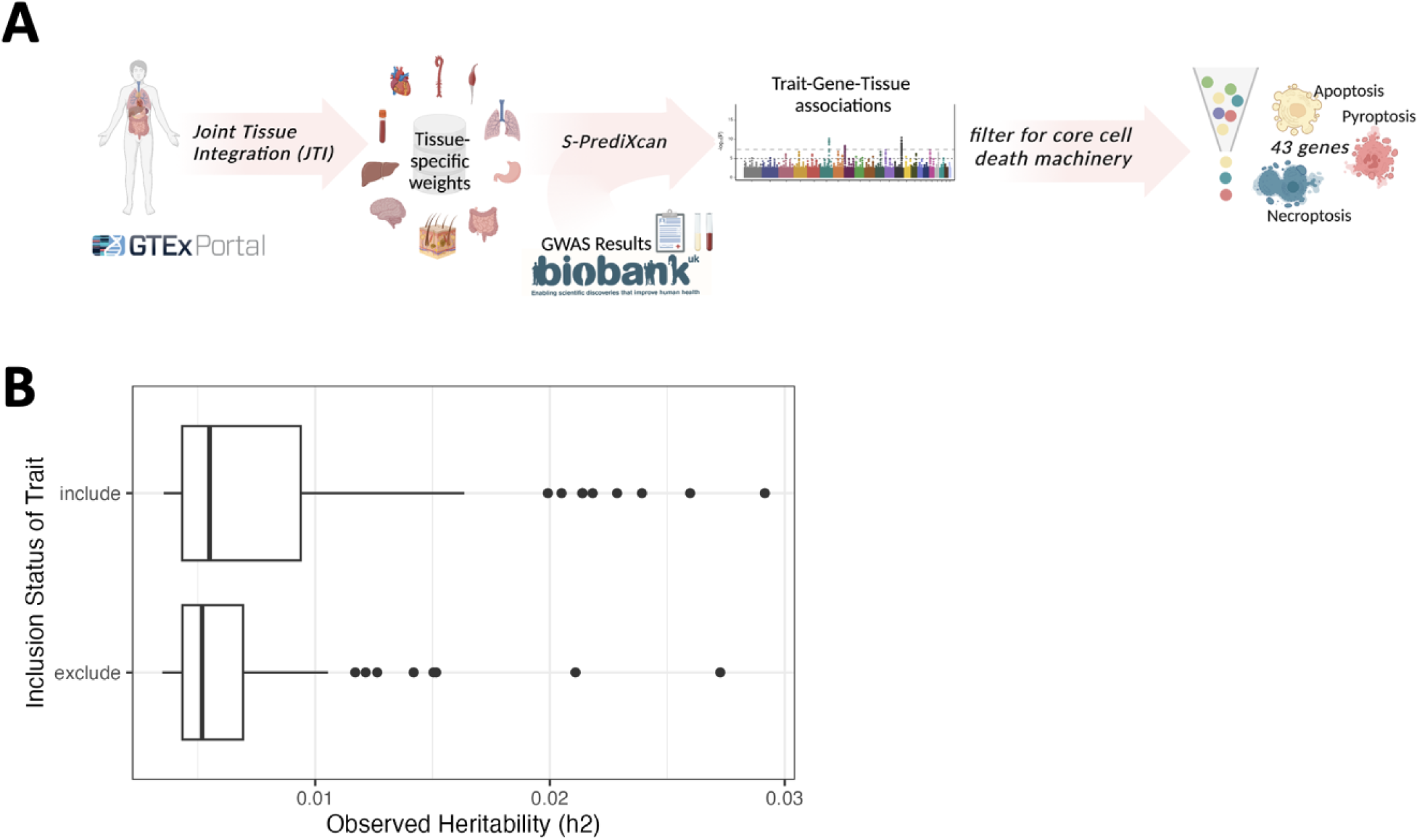
Joint-Tissue Integration and BioBank TWAS pipeline for phenome-wide scanning of cell death genes. 3A: Joint Tissue Integration was used to generate tissue-specific models of gene expression using the GTEx v8 resource, and these models were applied to both clinical diagnoses and lab-derived traits from the UKBB, which generated a series of gene-trait-tissue associations that enabled analysis of 43 cell death-associated genes. 3B: Observed heritability for clinical outcome traits that were included or excluded from analysis per our manual curation strategy.

#### Manual curation of relevant clinical outcome and lab-derived traits from the UKBB enriches for heritable traits

To maximize the power of our analysis and enhance the interpretability of findings, we manually curated a list of clinically relevant traits from the UKBB version 3 release (Supplemental Data). This curation prioritized quantitative traits from lab-derived tests and electronic health record (EHR)-derived clinical outcome traits with clear diagnoses, specific etiology, and/or minimal redundancy with other UKBB GWAS entries. Ultimately, we selected 482 EHR-derived clinical outcome traits from the UKBB and 31 lab-derived clinical traits from quantitative blood and urine measurements for analysis (Supplemental Table 6). The clinical outcome traits span 16 discrete phenotypic categories, and the 24 lab-derived traits encompass both blood and urine-derived markers (Supplemental Table 6). The average heritability of selected clinical outcome traits (as determined by linkage disequilibrium score regression; Supplemental Data) was higher than that of omitted clinical outcome traits (Fig. 3B). This suggests that our filtering strategy not only enriches for traits with clinical relevance, but traits for which these genetically informed analyses are more relevant.

The continuous lab-derived traits and binary clinical outcome traits had marked differences in sample size and the number of significant associations (Supplemental Fig. 2D). As such, lab traits were analyzed separately from clinical traits. Associations from the 24 lab-derived traits were analyzed with an FDR cutoff of 0.01. Given the phenome-wide scope of the EHR-derived trait analysis and subsequent multiple testing burden, to maximize the discovery potential for EHR-derived trait associations, we used an FDR cutoff of 0.25 (corresponding with an unadjusted p-value 7.7e-5).

### Genetically determined expression of cell death genes associates with health-related traits across the human phenome, and is unique across specific genes and traits

For EHR-derived traits, we identified 157 significant associations in 21 cell death genes across 27 unique health-related traits and 12 phenotype categories (Supplemental Table 7). Significant associations were detected across all 49 examined tissues (Supplemental Table 7). The number of significant associations identified for a given cell death gene/tissue pairing was not correlated with the number of tissue samples used to generate the tissue-specific models (i.e., weights), suggesting that tissue biology rather than sample size drives the number of associations (Supp. Fig. 3A). Similarly, the number of significant associations detected was not correlated with the number of cases used to perform association studies on clinical traits, suggesting that these analyses can detect *bona fide* biological signals (Supp. Fig. 3B). Though tissue specificity is an important biological consideration, we found that for gene/trait associations in multiple tissues where p<0.05, the overwhelming majority of associations were concordant in their direction of effect (Supplemental Table 7). This phenomenon empowers us to interpret TWAS results independent of the tissue model from which a given association arose and implies that, for traits with multiple tissue associations, there is a shared genetic architecture that drives associations across tissues.

We observed that genetically determined expression of a suite of cell death genes is associated with a range of clinically relevant traits comprising over a dozen different phenotypic categories (Fig. 4A). The most highly significant association was between *CASP8* expression and “Other malignant neoplasms of skin”. *CASP8* associations could be detected in a variety of different tissues (Supplemental Table 7, Fig. 4A). This suggests that there are tissue-shared eQTLs for the gene driving the associations across tissues. Significant associations were observed in genes involved in all examined cell death pathways, involving apoptotic, necroptotic, and pyroptotic genes (Fig. 4A). These findings reveal a potential link between genetically regulated expression of apoptosis, necroptosis, and pyroptosis machinery in the etiology of a variety of clinically relevant disorders.

**FIGURE 4:**
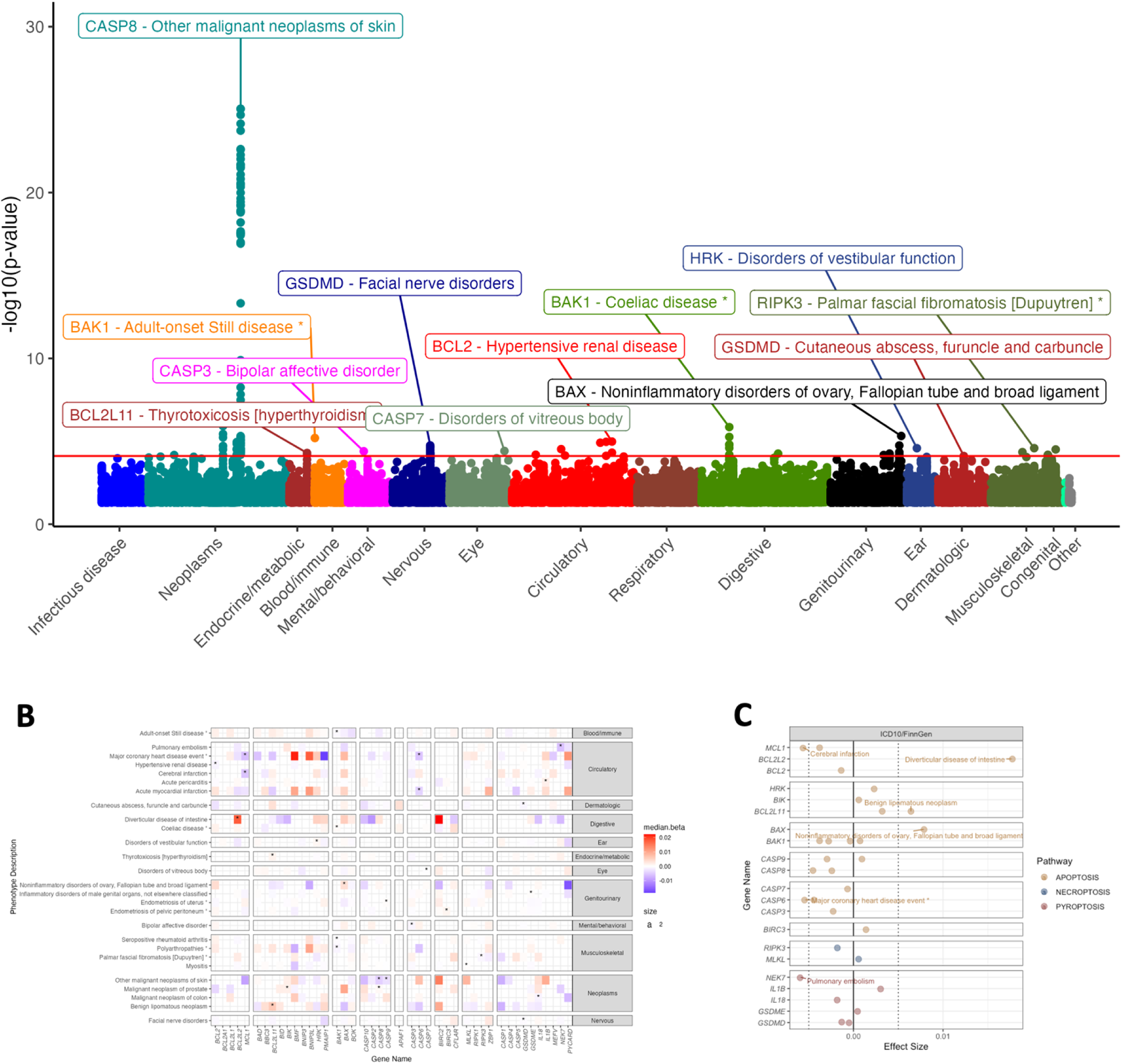
UKBB clinical diagnoses associated with genetically regulated expression of cell death genes. 4A: Manhattan plot illustrating top gene-trait associations by p-value and organized by trait type. Only the most significant gene-trait association in a phenotype category is labeled. The red line indicates the FDR=0.25 threshold (p_unadj_< 7.7e-5). Gene-trait associations with significant associations across multiple tissues (e.g., *CASP8* and Other malignant neoplasms of skin), are not annotated for clarity. 4B: Heatplot illustrating, for traits with n≥1 associations with FDR<0.25, median effect size of associations for said trait with p<0.05. Gene-trait associations with FDR<0.25 indicated by “*”. 4C: Median effect size for significant (FDR<0.25) gene/trait/tissue associations where associations with |β|>0.005 are labeled.

A unique feature of the gene-level TWAS approach is its ability to estimate both effect size and direction of effect for associations. Given the conserved structural and functional characteristics of many of the genes in our cell death array, we sought to resolve significant gene-trait association effect sizes by both gene similarity and trait similarity (Fig. 4B). We were surprised that not more significant associations occurred in groups of genes with functional similarities (e.g., Initiator Caspases, prosurvival BCL-2 family genes, as grouped on the X-axis in Fig. 4B). To enrich for potential additional relevant patterns, we graphed median effect sizes for nominally significant (p_unadj_<0.05) gene/trait associations alongside the significant gene/trait associations (as denoted by asterisks, Fig. 4B). The effect size and direction of shared associations for a particular trait was variable (Fig. 4B). Across significant (FDR<0.25) gene/trait associations, the largest magnitude of effect was the association of diverticular disease with lower expression of *BCL2L2*, encoding BCL-W (Fig. 4C). Explicitly, this means that the genetically regulated expression of BCL-W accounts for almost 3% of the genetic predisposition to diverticular disease of the intestine. These results provide preliminary evidence there is a directional relationship between cell death gene expression and a suite of human traits and support a model in which functionally similar genes and pathways have distinct roles in the etiology of disease.

### Laboratory traits are enriched for significant associations with hematologic phenotypes

We identified hundreds of significant (FDR<0.01) gene/tissue/trait associations for lab-derived traits (Supplemental Table 8). The most highly significant associations were in blood traits, particularly associations between *BAK1* and platelet count & crit (Table 3). Other highly significant associations were observed between leukocyte blood parameters and *MCL1*, and *BCL2L1* (encoding BCL-XL) and red blood cell parameters (Fig. 5A). As was observed in our analysis of EHR-derived traits, the direction of effect for gene/trait/tissue associations across tissues were overwhelmingly concordant for any given trait (Supplemental Table 8).

**FIGURE 5:**
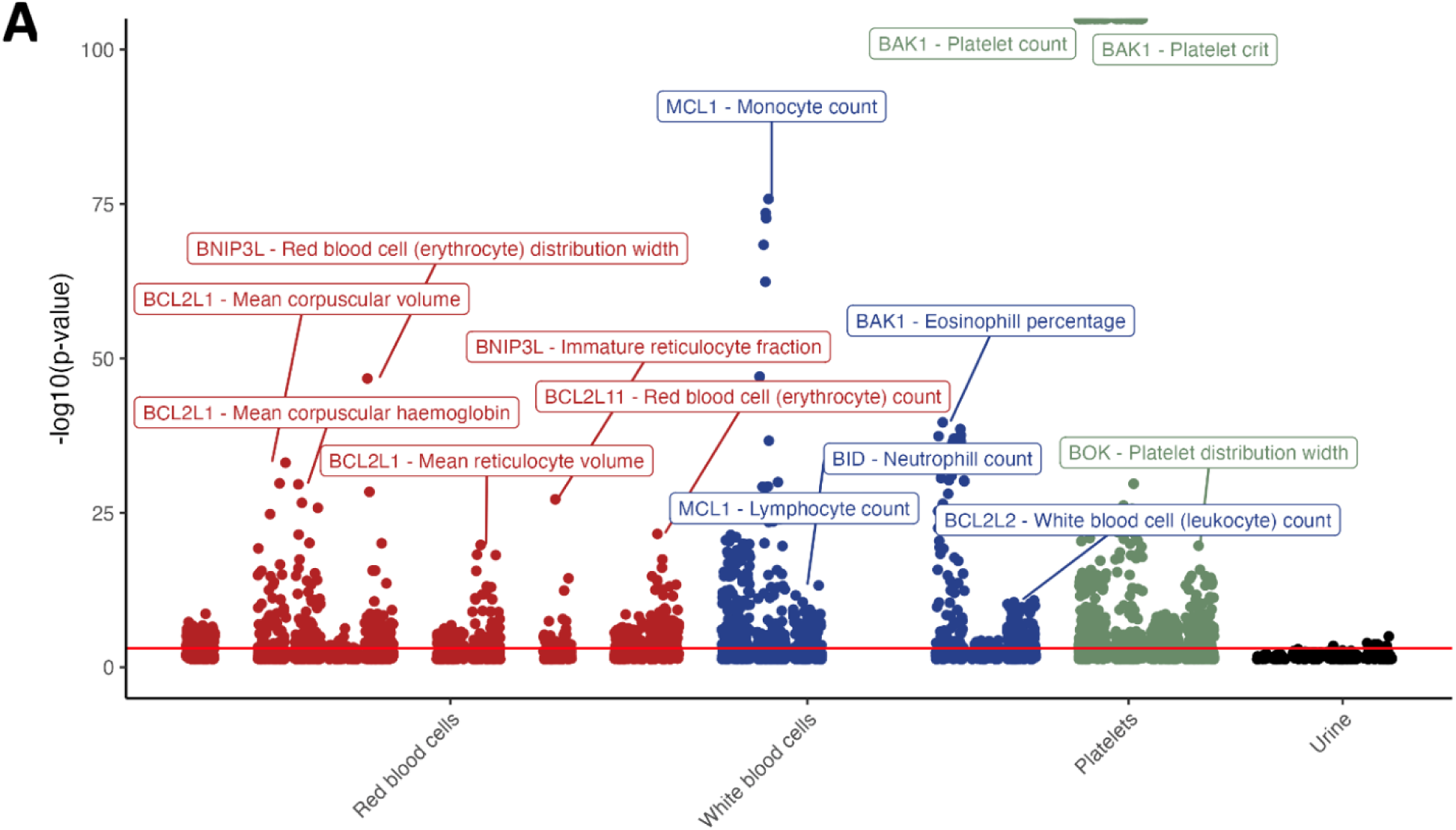
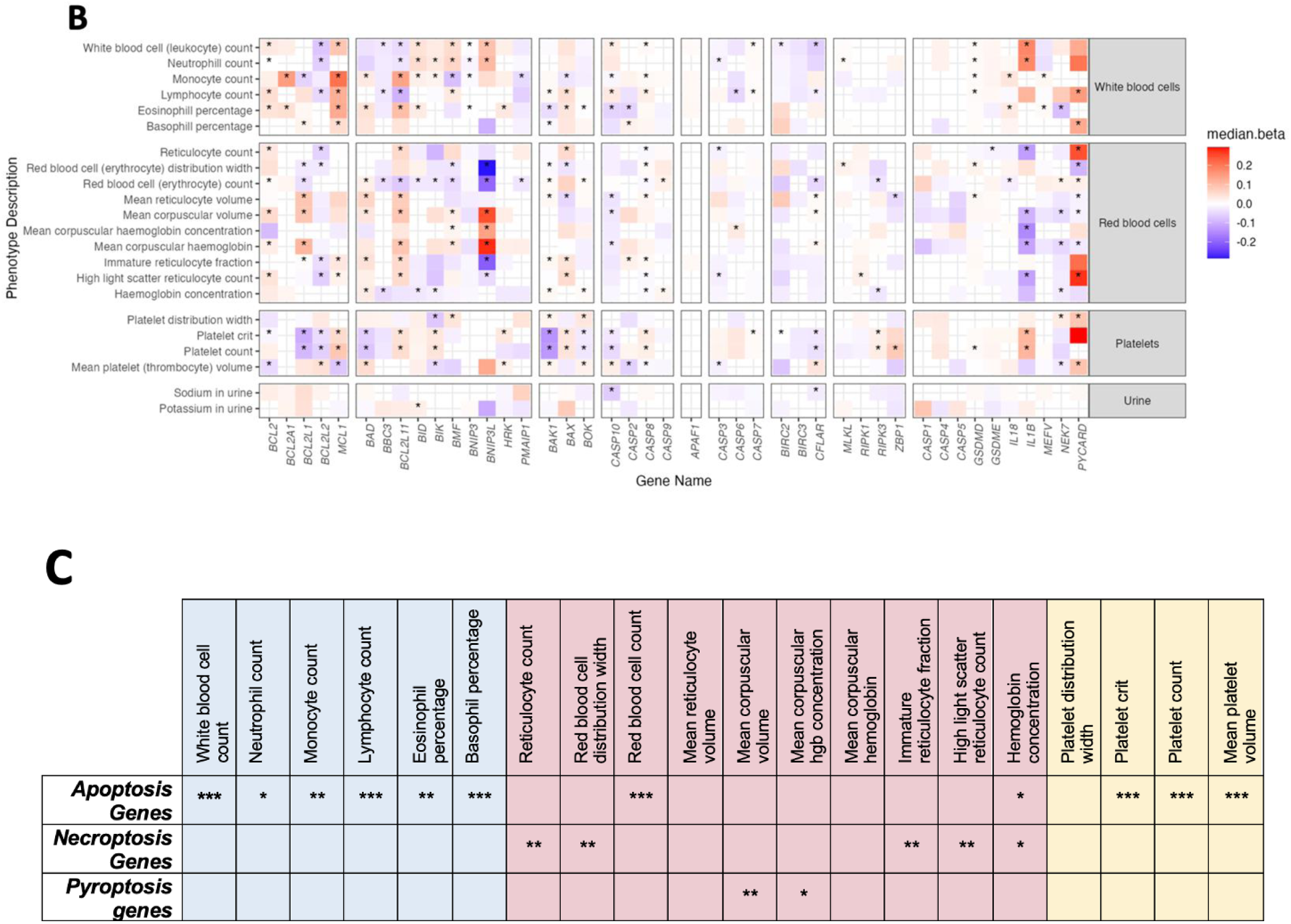
Lab-derived continuous traits associated with genetically regulated expression of cell death genes. 5A: Manhattan plot illustrating top laboratory-derived gene-trait associations by p-value and organized by trait type. Unique trait associations with p_unadj_<2e-10 are labeled, and the red line illustrates the FDR=0.01 threshold (p<0.0008353). Gene/trait associations across multiple tissues are omitted from this plot for clarity. 5B: Median effect size and direction across all gene/trait pairs with a tissue association of p<0.05. Significant (FDR<0.01) gene/trait associations are indicated by an asterisk. 5C: Fisher’s Exact testing for enrichment of associations for apoptosis, necroptosis, and pyroptosis genes as defined in Supplemental Table 1. *p<0.05, **p<0.01, ***p<5e-10.

Resolving significant (FDR<0.01) and nominally significant (p<0.05) association effect sizes by gene family and trait family reveal a matrix of gene/trait associations that are not redundant across similar gene families (Fig. 5B; e.g., *BCL2L1* encoding BCL-XL and *MCL1* display associations with opposite directions of effect across blood traits). The most extreme associations by median effect size were observed in red blood cell-associated traits and with BH3-only gene *BNIP3L* and pyroptosis-associated adapter *PYCARD* (Fig. 5B). For many of these associations, up to 20% of the genetic contribution for several traits can be traced back to genetically determined expression of one of these two genes (Fig. 5B, Supplemental Table 8). On the whole, these findings reinforce the proposition there are distinct roles for cell death genes that are considered functionally redundant in their contribution to human phenotypes and indicate that the genetically determined expression of *BNIP3L* and *PYCARD* exert a significant effect on hematologic, particularly red blood cell traits.

There were many more highly significant associations with large-magnitude effect sizes for blood cell traits relative to urine traits (Fig. 5A-B), suggesting that cell death gene expression is uniquely important for shaping hematopoiesis. To explicitly test if cell death pathway genes were overrepresented among our significant associations, we performed enrichment analysis on apoptotic, necroptotic, or pyroptotic gene sets across each of the blood traits. Cell death genes were highly enriched for significant associations with blood traits versus non-cell-death genes, with apoptosis gene sets enriched for significant associations with leukocyte and platelet traits, and necroptosis and pyroptosis gene sets enriched for associations with erythroid traits (Fig. 5C). Overall, these results suggest that the genetically regulated expression of cell death genes are particularly important in shaping the numbers and distributions of blood and immune cells.

### Replication of gene/trait associations in an independent large-scale biobank

Our discovery analysis implicated genetically determined expression of cell death genes in dozens of clinically-relevant diagnoses that are subject to replication. We performed a limited external replication analysis by applying our TWAS methodology to BioVU^48^. We prioritized top gene/trait associations that had corresponding phecodes in the BioVU dataset. Remarkably, we detected significant associations between *BAK1* and Rheumatoid arthritis/Polyarthropathies (p=2.50e-3) as well as *BCL2L2* and Diverticular disease of the intestine (p=3.86e-02) (Table 1). This replication validates at least three associations (for Rheumatoid arthritis, polyarthropathies, diverticulitis) found within our study and support the utility of our approach to discovering novel gene/trait associations.

**Table 1.** Independent replication analysis in the BioVU biobank.

| phecode | effect | p-value | gene | phenotype | # cases | # controls |
| --- | --- | --- | --- | --- | --- | --- |
| **714** | -0.282262 | 2.50E-03 | BAK1 | Rheumatoid arthritis and other inflammatory polyarthropathies | 928 | 19052 |
| **562.2** | 0.800272 | 3.86E-02 | BCL2L2 | Diverticulitis | 291 | 16064 |

## DISCUSSION

In this study, we present a novel transcriptomic survey of cross-tissue cell death gene expression and deliver an atlas of human traits that are influenced by genetically determined expression of core apoptotic, necroptotic, and pyroptotic genes. Our transcriptomic analyses reveal there is tissue-specific patterning in the expression of cell death pathway genes and identify a diverse array of clinically relevant traits that are associated with genetically regulated expression of cell death genes. Our work implies that dysregulation of cell death gene expression can contribute to a wide variety of human phenotypes, suggests that functionally similar cell death gene paralogs have unique contributions to human pathophysiology, and supports a paradigm in which cell death pathways operate heterogeneously across tissues.

Our analysis identified dozens of human phenotypes that are associated with cell death gene expression, both novel and previously reported. Among novel gene/trait associations of potential interest to the cell death field, we identify an association between increased *BCL2L2* (encoding BCL-W) and its association with diverticular disease of the intestine and *CASP3* and bipolar affective disorder. Our findings align with previously identified cell death/human disease relationships. The most significant association was between *CASP8* and “Other malignant neoplasms of skin”. Indeed, *CASP8* mutations have been reported in cutaneous squamous cell carcinoma^49^. Interestingly, all initiator caspases (caspases -8, -9, and -10) demonstrated at least nominally significant, unidirectional associations with malignant skin neoplasms (Fig. 4B), suggesting that reduced expression of any initiator caspases may significantly promote skin malignancy. A contingent of superinflammatory disorders were associated increased expression of cell death genes participating in immunogenic cell death (*IL1B*/acute pericarditis, *MLKL*/myositis, *GSDME*/inflammatory disorders of male genital organs), meanwhile, decreased expression of the proapoptotic effector gene *BAK1* associated with the proinflammatory disorders, adult onset Stills disease and seropositive rheumatoid arthritis.

Observations from mouse models (in which germline and/or conditional knockouts serve as a proxy for extremely low gene expression) also support the validity of our findings as well. We observed that lower expression of BNIP3L associates with high RBC and reticulocyte counts (Fig. 5B). In mice, knockout of *Bnip3l* (mouse ortholog of *BNIP3L*) induces profound erythrocytosis^50^. *Mcl1* knockout mice develop cardiac insufficiencies that culminate in heart failure^51, 52^; our PheWAS analysis identified a significant association between low *MCL1* expression and “Major coronary heart disease event”.

TWAS analyses such as those that form the basis of our PheWAS have utility in their ability to identify relationships between extremes in gene expression and traits. Though associations do not imply causal relationships (i.e., lower expression of *GeneX* causes *DiseaseY*), the findings may be of utility in the vetting and application of cell death inhibitory compound administration, as the outcome of restricted gene expression is lower protein content/function. Indeed, gene expression values and protein levels as determined by mass spectrometry (Human Protein Atlas dataset reference) were significantly positively correlated (Supp. Fig. 2A), suggesting that gene expression analyses may extrapolate to protein-level phenomena. We observed that low expression of *BCL2L1* (encoding BCL-XL) is significantly associated with a decrease in platelet count and platelet crit (Fig. 5B). Correspondingly, the major dose-limiting toxicity of an inhibitor of BCL-XL, navitoclax, is its reduction in platelet count^53^. Many of the tested genes have associations with multiple hematopoietic traits, highlighting the highly interdependent nature of hematopoietic cell differentiation and the importance of cell death for hematopoiesis. For instance, increased expression of *BCL2L1* is strongly positively correlated with mean corpuscular volume (MCV) and mean reticulocyte volume (Fig. 5B). This is consistent with observations that BCL-XL deficiency impairs late erythroblast/reticulocyte survival^54^.

An intriguing phenomenon across functionally similar gene groupings (defined in Fig. 1) was the presence of significant gene/trait associations with opposite directions of effect. For example, *BAX* and *BAK1* are thought to be functionally redundant, as dual knockout of these pore-forming proteins is required to elicit multi-organ pathologies^55^. *BAK1* and *BAX* had divergent median betas when considering platelet count/crit, eosinophil percentage, and mean reticulocyte volume (Fig. 5B). Furthermore, *BCL2L1* and *MCL1*, both prosurvival proteins, had discordant associations with platelet count & platelet crit, monocyte count, and immature reticulocyte fraction (Fig. 5B). These results highlight the potential for non-redundant roles of apoptotic family genes in human hematologic traits.

Though there are gene-specific patterns across all gene/trait associations from our analyses, overall, it is clear that cell death gene expression associates with a multitude of hematologic traits. Cell death genes were highly enriched for significant associations with blood traits versus non-cell-death genes, with apoptosis-associated genes enriched for leukocyte and platelet traits and necroptosis gene associations enriched for erythroid traits (e.g., Reticulocyte count, FDR=0.004) (Fig. 5C). This suggests that immunogenic/proinflammatory cell death pathways play an important role in regulating erythropoiesis and reinforces the paradigm that apoptosis pathway genes are critical for white blood cell and platelet development.

Our cross-tissue survey of expression patterns revealed discrete tissue modules with correlated gene expression signatures (Fig. 2C). Using only our array of 44 genes, we identified biologically coherent tissue modules that segregated into relevant groupings: nervous tissues (module 1), which are comprised largely of a pool of post-mitotic cells, segregated clearly from a group of rapidly dividing tissues including intestines, whole blood, and skin (module 3), and these were separated by an intermediate module (2) with various somatic tissues. The most highly correlated gene pairs within each module (Fig. 2D-F) were surprisingly not gene paralogs, nor did many highly correlated genes reside in the same pathways, suggesting that cell death gene expression networks have unique architecture in disparate tissues (Supplemental Table 3). Notably, EBV-transformed lymphocytes comprised a module distinct from all primary tissues examined (Fig. 2C), suggesting that this cell type is divergent from other primary tissue types with regards to cell death gene expression. The unique transcriptional signature in these immortalized cells highlights a limitation of using transformed lymphocytes in studies in which the dynamics of cell death are important for the readout (e.g., drug toxicity screens, MPRAs). Indeed, the EBV-encoded E1B protein functionally complements BCL-2 in its anti-apoptotic action to facilitate lymphocyte transformation^56^. As such, these findings advocate for careful selection of mechanistic models that avoid a non-immortalized cell lines and recapitulate the transcriptional “footprint” of the target tissue or organ system.

This study has limitations that must be considered in the interpretation of our findings. The TWAS methodology predicts how germline, rather than somatically acquired, genetic variants influence expression of a gene based on a reference transcriptome dataset. The statistical models developed for our analyses rely upon training data acquired from adult somatic tissues (GTEx v8). As such, these analyses cannot make conclusions about how acquired genetic mutations (e.g., BCL2:IGH translocations) influence human phenotypes. Rare gain- or loss-of function mutations in our cell death gene array, that may have a substantial impact on the functioning of these pathways, are also not captured in this study. Such variants have been captured by Karczewski and colleagues (2022)^57^, who identified germline rare variants in cell death genes that associate with human traits via whole exome sequencing of large populations. Databases for interrogating pathogenic mutations of cell death genes and protein structures/post-translational modifications are described more deeply elsewhere^58^. It follows that our approach cannot discriminate between adult and developmental expression patterning in the etiology of disorders. The clustering of observed expression data and directionality of gene/trait associations transcend classical cell death pathways and may support emerging literature implicating the extensive crosstalk between these pathways^59, 60^. Our enrichment analyses as presented do not consider gene/trait signatures in the context of crosstalk signaling nodes, which our data suggest may contain relevant and exciting biological signals.

Our transcriptome training models were derived from the sampled individuals in GTEx v8, and the GWAS summary statistics data were derived from the UKBB, resources that are biased towards individuals of European ancestry, limiting our ability to make cross-ancestry generalizations. Some disorders have an independently and rigorously established link to defects in the expression of cell death genes, such as *CASP8* and *CASP10* and autoimmune lymphoproliferative syndrome^33^ yet were not detected by our analysis. The number of cases in the UKBB v3 release for some disorders was small (Supplemental Data) and as such could have limited our power to detect associations with these disorders. The specificity of diagnoses impact both case counts and one’s ability to interpret associations (for instance “Other malignant neoplasms of skin” diagnosis code may encompass a range of distinct etiologies and does not correspond directly to a single phenotype designation in our replication dataset).

Our study identified and replicated novel associations between genetically determined expression of cell death genes and human traits, defined hundreds of significant (FDR<0.01) associations with lab derived traits, and found blood traits significantly enriched for associations with specific cell death pathways. These findings can be further replicated in other large-scale biobanks. Overall, these results suggest that the genetically regulated expression of cell death genes are particularly important in shaping the numbers and distributions of blood and immune cells, reinforce the proposition there are *distinct* roles for cell death genes that are classically considered functionally redundant in their contribution to human phenotypes, and emphasize that cell death genes are involved in the determination of human traits across the phenome.

## METHODS

### Parameters for Selection of Cell Death Gene Array

We focused on apoptotic, necroptotic and pyroptotic signaling pathways to conscribe the bounds of the analysis. Intrinsic and extrinsic apoptosis, necroptosis, and pyroptosis are examples of highly studied and well-defined pathways whose genetically encoded machinery participates primarily in the process of cell death, with minimal shunting to metabolic pathways. Omitted were cell surface receptors initiating these pathways, enzymes involved in non-core post-translational modifications of pathway members, and pathway proteins that participate in but operate on the periphery of these pathways (for instance, *FADS1/2* which are important for ferroptosis also participate in fatty acid metabolism and contribute to other aspects of cell function & fate).

### Gene Expression Correlation & Clustering Analyses

Preprocessed GTEx v8 tissue expression data in transcripts per million (TPM) from 49 tissues were extracted for analysis. Median TPM values for each gene in the of the cell death gene array were calculated each tissue, and then applied to Pearson correlation analysis. Ward’s D hierarchical clustering was implemented to identify four discrete tissue modules by graphing the correlation coefficients using the ‘corrplot’ v0.92 package in R 4.2.1. Euclidean distance between median gene expression TPMs across modules was applied to generate dendrograms for gene-gene relationships across modules.

### Manual Curation of Health-Related Outcome Traits

To enrich our results for clinically informative traits, we considered health-related outcome phenotypes derived from ICD10 Diagnosis codes (UK Biobank Data Field 41270) as well as multi-parameter phenotypes derived in collaboration with the FinnGen consortium (“FinnGen custom” phenotypes) from the UKBB GWAS round 2 analysis (see Supplemental Data). These encompass 1144 unique phenotypes (Supplemental Table 6) with varying degrees of overlap. ICD10 codes categorized as representing “Pregnancy, childbirth, and the pueperium” (Chapter XV / “O” prefix), “Symptoms, signs and abnormal clinical and laboratory findings, not otherwise classified” (Chapter XVIII / “R” prefix), “Injury poisoning and certain other consequences of external causes” (Chapter XIX / “S” and “T” prefixes), and “Factors influencing health status and contact with health services” (Chapter XXI / “Z” prefixes) were excluded from analysis, as they generally have lower genetic heritability as estimated by linkage disequilibrium score regression (Fig. 3B). GWAS summary statistics calculated after sex stratification were omitted from our analysis as well. Additional manual review of these traits removed FinnGen custom phenotypes that overlapped with more specifically defined or identical ICD10 Diagnosis Codes as well as phenotypes representing nonspecific or “catch-all” traits (Supplemental Data).

Laboratory-derived traits with measurements derived from quantitative assays were selected for analysis, and GWAS summary statistics that were calculated using inverse rank-based normal transformation (IRNT) were chosen for final analysis. This resulted in 31 continuous traits derived from blood and urine tests. Of these traits, some measurements derived from complete blood count data were redundant and removed. These were: monocyte percentage, lymphocyte percentage, neutrophil percentage, high light scatter reticulocyte percentage, reticulocyte percentage, and mean sphered cell volume.

### Joint Tissue Imputation (JTI) and Transcriptome-Wide Association Study (TWAS)

The summary statistic-based S-PrediXcan methodology developed by Barbiera *et al*. (2018) was employed using tissue-specific models generated using the Joint Tissue Imputation (JTI) methodology developed by Zhou *et al*. (2020).

Overall, 1,061 gene/tissue pairs x 513 traits were assessed, resulting in 544,293 individual tests. The Benjamini-Hochberg multiple hypothesis testing correction was employed to adjust for this large multiple testing burden. A false-discovery rate cutoff of 25% was chosen for clinical outcomes traits to enable a trait discovery-based analysis. Given the higher sample size and sensitivity of continuous trait analysis, we opted to use a more stringent FDR cutoff of 1% for data presentation and enrichment analysis.

### Replication analysis

We performed limited replication testing of our most significant findings (Table 1) in an independent biobank linked to EHR data, Vanderbilt’s BioVU repository. We tested *BAK1*’s association with Rheumatoid arthritis and with polyarthropathies, *BCL2L2*’s association with Diverticulitis, and *MCL1*’s association with cerebral infarction BioVU. We were unable to test for replication of *CASP8*’s association with “Other malignant neoplasms of skin,” our most significant association, due to lack of a single phenotype designation in the replication dataset.

## DATA AVAILABILITY

JTI was performed using GTEx v8 transcriptomic data (dbGaP phs000424.vN.pN). GWAS summary statistics and heritability data were obtained from the publicly-available UKBB v3 release at http://www.nealelab.is/uk-biobank and https://nealelab.github.io/UKBB_ldsc/downloads.html, respectively. Source code and implementation notes for S-PrediXcan and JTI, as well as custom code for figure generation and analyses can be found in this GitHub repository (to be made public at the time of publication).

## Data Availability

JTI was performed using GTEx v8 transcriptomic data (dbGaP phs000424.vN.pN). GWAS summary statistics and heritability data were obtained from the publicly-available UKBB v3 release at http://www.nealelab.is/uk-biobank and https://nealelab.github.io/UKBB_ldsc/downloads.html, respectively.

## ACKNOWLEDGEMENTS

We acknowledge support from the following National Institutes of Health (NIH) grants: NHGRI R35HG010718, NHGRI R01HG011138, NIMH R01MH126459, NIGMS R01GM140287, NIA AG068026, NHLBI R01HL133559, and VA MERIT 5I01BX004365.

## AUTHOR INFORMATION

These authors wrote the manuscript: Abigail L. Rich, Sandra S. Zinkel, and Eric R. Gamazon.

These authors performed analyses: Abigail L. Rich & Phillip Lin.

These authors jointly supervised this work: Sandra S. Zinkel & Eric R. Gamazon.

## REFERENCES

1. Kerr, J. F. R., Wyllie, A. H. & Currie, A. R. Apoptosis: A Basic Biological Phenomenon with Wide-ranging Implications in Tissue Kinetics. Br J Cancer 26, 239–257 (1972).

2. Vaux, D. L. & Korsmeyer, S. J. Cell Death in Development. Cell 96, 245–254 (1999).

3. Green, D. R. Cell Death in Development. Cold Spring Harb Perspect Biol 14, a041095 (2022).

4. Tower, J. Programmed cell death in aging. Ageing Research Reviews 23, 90–100 (2015).

5. Ketelut-Carneiro, N. & Fitzgerald, K. A. Apoptosis, Pyroptosis, and Necroptosis—Oh My! The Many Ways a Cell Can Die. Journal of Molecular Biology 434, 167378 (2022).

6. Danial, N. N. & Korsmeyer, S. J. Cell Death: Critical Control Points. Cell 116, 205–219 (2004).

7. Strasser, A., O’Connor, L. & Dixit, V. M. Apoptosis signaling. Annu Rev Biochem 69, 217–245 (2000).

8. Linkermann, A. & Green, D. R. Necroptosis. N Engl J Med 370, 455–465 (2014).

9. Zhou, W. & Yuan, J. Necroptosis in health and diseases. Seminars in Cell & Developmental Biology 35, 14–23 (2014).

10. Broz, P. & Dixit, V. M. Inflammasomes: mechanism of assembly, regulation and signalling. Nat Rev Immunol 16, 407–420 (2016).

11. Weinlich, R., Oberst, A., Beere, H. M. & Green, D. R. Necroptosis in development, inflammation and disease. Nat Rev Mol Cell Biol 18, 127–136 (2017).

12. Khoury, M. K., Gupta, K., Franco, S. R. & Liu, B. Necroptosis in the Pathophysiology of Disease. The American Journal of Pathology 190, 272–285 (2020).

13. Bertheloot, D., Latz, E. & Franklin, B. S. Necroptosis, pyroptosis and apoptosis: an intricate game of cell death. Cell Mol Immunol 18, 1106–1121 (2021).

14. Andreone, B. J., Larhammar, M. & Lewcock, J. W. Cell Death and Neurodegeneration. Cold Spring Harb Perspect Biol 12, a036434 (2020).

15. Basiorka, A. A. et al. The NLRP3 inflammasome functions as a driver of the myelodysplastic syndrome phenotype. Blood 128, 2960–2975 (2016).

16. Wagner, P. N. et al. Increased Ripk1-mediated bone marrow necroptosis leads to myelodysplasia and bone marrow failure in mice. Blood 133, 107–120 (2019).

17. Zou, J., Shi, Q., Chen, H., Juskevicius, R. & Zinkel, S. S. Programmed necroptosis is upregulated in low-grade myelodysplastic syndromes and may play a role in the pathogenesis. Experimental Hematology (2021) doi:10.1016/j.exphem.2021.09.004.

18. Hanahan, D. & Weinberg, R. A. The Hallmarks of Cancer. Cell 100, 57–70 (2000).

19. Hanahan, D. & Weinberg, R. A. Hallmarks of Cancer: The Next Generation. Cell 144, 646–674 (2011).

20. Hanahan, D. Hallmarks of Cancer: New Dimensions. Cancer Discovery 12, 31–46 (2022).

21. Zinkel, S. S. et al. Proapoptotic BID is required for myeloid homeostasis and tumor suppression. Genes Dev. 17, 229–239 (2003).

22. Ranger, A. M. et al. Bad-deficient mice develop diffuse large B cell lymphoma. Proceedings of the National Academy of Sciences 100, 9324–9329 (2003).

23. Cory, S., Vaux, D. L., Strasser, A., Harris, A. W. & Adams, J. M. Insights from Bcl-2 and Myc: malignancy involves abrogation of apoptosis as well as sustained proliferation. Cancer Res 59, 1685s–1692s (1999).

24. Pezzano, M. et al. Positive Selection by Thymic Nurse Cells Requires IL-1β and Is Associated with an Increased Bcl-2 Expression. Cellular Immunology 169, 174–184 (1996).

25. Campbell, K. J., Gray, D. H. D., Anstee, N., Strasser, A. & Cory, S. Elevated Mcl-1 inhibits thymocyte apoptosis and alters thymic selection. Cell Death Differ 19, 1962–1971 (2012).

26. Mandal, M. et al. Regulation of lymphocyte progenitor survival by the proapoptotic activities of Bim and Bid. Proceedings of the National Academy of Sciences 105, 20840–20845 (2008).

27. Bouillet, P. et al. BH3-only Bcl-2 family member Bim is required for apoptosis of autoreactive thymocytes. Nature 415, 922–926 (2002).

28. Enders, A. et al. Loss of the Pro-Apoptotic BH3-only Bcl-2 Family Member Bim Inhibits BCR Stimulation– induced Apoptosis and Deletion of Autoreactive B Cells. Journal of Experimental Medicine 198, 1119–1126 (2003).

29. Mountz, J. D., Zhou, T., Su, X., Wu, J. & Cheng, J. The Role of Programmed Cell Death as an Emerging New Concept for the Pathogenesis of Autoimmune Diseases. Clinical Immunology and Immunopathology 80, S2–S14 (1996).

30. Takeuchi, O. et al. Essential role of BAX,BAK in B cell homeostasis and prevention of autoimmune disease. Proceedings of the National Academy of Sciences 102, 11272–11277 (2005).

31. Tischner, D., Woess, C., Ottina, E. & Villunger, A. Bcl-2-regulated cell death signalling in the prevention of autoimmunity. Cell Death Dis 1, e48–e48 (2010).

32. Casamayor-Polo, L. et al. Immunologic evaluation and genetic defects of apoptosis in patients with autoimmune lymphoproliferative syndrome (ALPS). Critical Reviews in Clinical Laboratory Sciences 58, 253–274 (2021).

33. Rieux-Laucat, F. What’s up in the ALPS. Current Opinion in Immunology 49, 79–86 (2017).

34. Sarosiek, K. A. et al. Developmental Regulation of Mitochondrial Apoptosis by c-Myc Governs Age- and Tissue-Specific Sensitivity to Cancer Therapeutics. Cancer Cell 31, 142–156 (2017).

35. Pellettieri, J. & Sánchez Alvarado, A. Cell turnover and adult tissue homeostasis: from humans to planarians. Annu Rev Genet 41, 83–105 (2007).

36. Barkett, M. & Gilmore, T. D. Control of apoptosis by Rel/NF-κB transcription factors. Oncogene 18, 6910– 6924 (1999).

37. Fridman, J. S. & Lowe, S. W. Control of apoptosis by p53. Oncogene 22, 9030–9040 (2003).

38. Molnár, T. et al. Current translational potential and underlying molecular mechanisms of necroptosis. Cell Death Dis 10, 1–21 (2019).

39. Aguet, F. et al. Genetic effects on gene expression across human tissues. Nature 550, 204–213 (2017).

40. Oleksiak, M. F., Churchill, G. A. & Crawford, D. L. Variation in gene expression within and among natural populations. Nat Genet 32, 261–266 (2002).

41. Barbeira, A. N. et al. Exploring the phenotypic consequences of tissue specific gene expression variation inferred from GWAS summary statistics. Nat Commun 9, 1825 (2018).

42. Green, D. R. The Coming Decade of Cell Death Research: Five Riddles. Cell 177, 1094–1107 (2019).

43. Cook, G. P., Savic, S., Wittmann, M. & McDermott, M. F. The NLRP3 inflammasome, a target for therapy in diverse disease states. European Journal of Immunology 40, 631–634 (2010).

44. Ashkenazi, A., Fairbrother, W. J., Leverson, J. D. & Souers, A. J. From basic apoptosis discoveries to advanced selective BCL-2 family inhibitors. Nat Rev Drug Discov 16, 273–284 (2017).

45. Martens, S., Hofmans, S., Declercq, W., Augustyns, K. & Vandenabeele, P. Inhibitors Targeting RIPK1/RIPK3: Old and New Drugs. Trends in Pharmacological Sciences 41, 209–224 (2020).

46. McIlwain, D. R., Berger, T. & Mak, T. W. Caspase Functions in Cell Death and Disease. Cold Spring Harb Perspect Biol 5, a008656 (2013).

47. Zhou, D. et al. A unified framework for joint-tissue transcriptome-wide association and Mendelian randomization analysis. Nat Genet 52, 1239–1246 (2020).

48. Denny, J. C. et al. PheWAS: demonstrating the feasibility of a phenome-wide scan to discover gene– disease associations. Bioinformatics 26, 1205–1210 (2010).

49. Chang, D. & Shain, A. H. The landscape of driver mutations in cutaneous squamous cell carcinoma. *npj Genom*. Med. 6, 1–10 (2021).

50. Diwan, A. et al. Unrestrained erythroblast development in Nix−/− mice reveals a mechanism for apoptotic modulation of erythropoiesis. Proceedings of the National Academy of Sciences 104, 6794–6799 (2007).

51. Thomas, R. L. et al. Loss of MCL-1 leads to impaired autophagy and rapid development of heart failure. Genes Dev. 27, 1365–1377 (2013).

52. Wang, X. et al. Deletion of MCL-1 causes lethal cardiac failure and mitochondrial dysfunction. Genes Dev. 27, 1351–1364 (2013).

53. Xiong, H. et al. Studying Navitoclax, a Targeted Anticancer Drug, in Healthy Volunteers – Ethical Considerations and Risk/Benefit Assessments and Management. Anticancer Research 34, 3739–3746 (2014).

54. Rhodes, M. M., Kopsombut, P., Bondurant, M. C., Price, J. O. & Koury, M. J. Bcl-xL prevents apoptosis of late-stage erythroblasts but does not mediate the antiapoptotic effect of erythropoietin. Blood 106, 1857– 1863 (2005).

55. Lindsten, T. et al. The Combined Functions of Proapoptotic Bcl-2 Family Members Bak and Bax Are Essential for Normal Development of Multiple Tissues. Molecular Cell 6, 1389–1399 (2000).

56. Rao, L. et al. The adenovirus E1A proteins induce apoptosis, which is inhibited by the E1B 19-kDa and Bcl-2 proteins. Proceedings of the National Academy of Sciences 89, 7742–7746 (1992).

57. Karczewski, K. J. et al. Systematic single-variant and gene-based association testing of thousands of phenotypes in 394,841 UK Biobank exomes. Cell Genomics 2, (2022).

58. Aouacheria, A., Navratil, V. & Combet, C. Database and Bioinformatic Analysis of BCL-2 Family Proteins and BH3-Only Proteins. in *BCL-2* Family Proteins: Methods and Protocols (ed. Gavathiotis, E.) 23–43 (Springer, 2019). doi:10.1007/978-1-4939-8861-7_2.

59. Frank, D. & Vince, J. E. Pyroptosis versus necroptosis: similarities, differences, and crosstalk. Cell Death Differ 26, 99–114 (2019).

60. Pandian, N. & Kanneganti, T.-D. PANoptosis: A Unique Innate Immune Inflammatory Cell Death Modality. The Journal of Immunology 209, 1625–1633 (2022).

