## Supplementary Figures for "The broad impact of cell death genes on the human disease phenome"

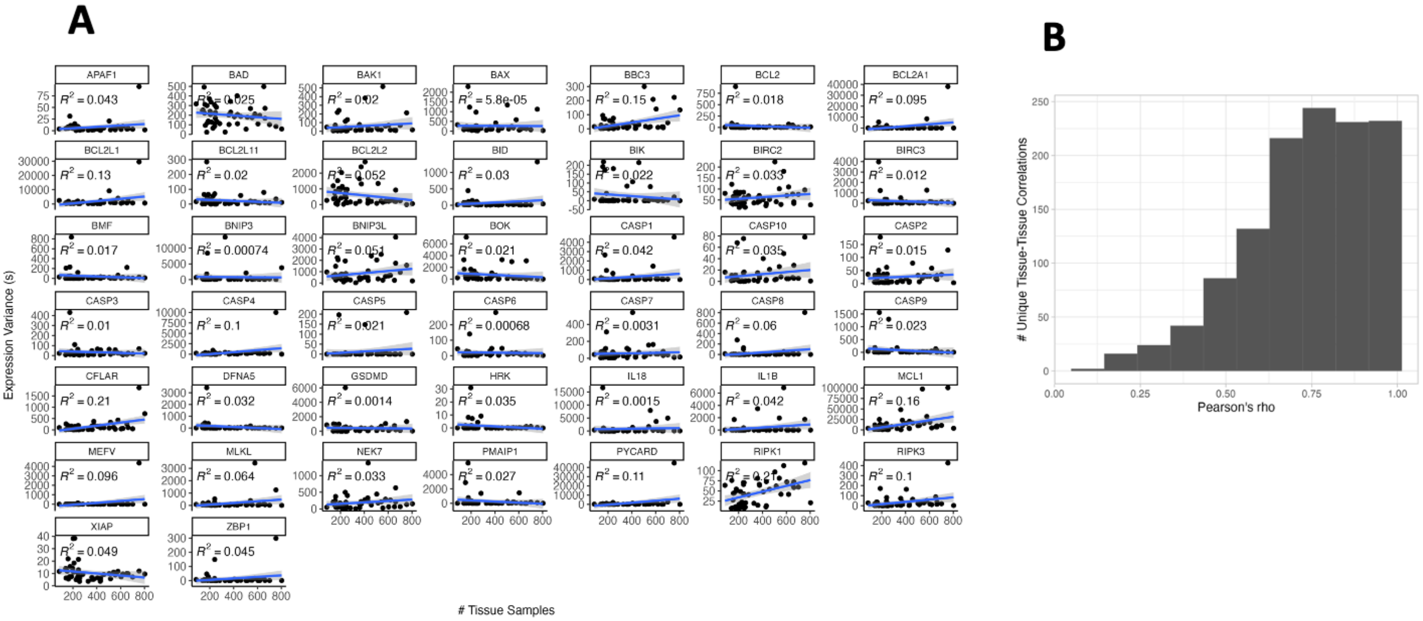


**Supplemental Figure 1: Observed cell death gene expression correlations.**

1A: Linear relationship between the number of GTEx v8 samples with gene expression data for a tissue and the variance in the sample's gene expression values (in TPM) across the cell death genes examined.

1B: Histogram depicting the frequency of tissue-tissue correlation values arising from correlation analysis of median TPM of cell death gene array transcripts in GTEx tissues.

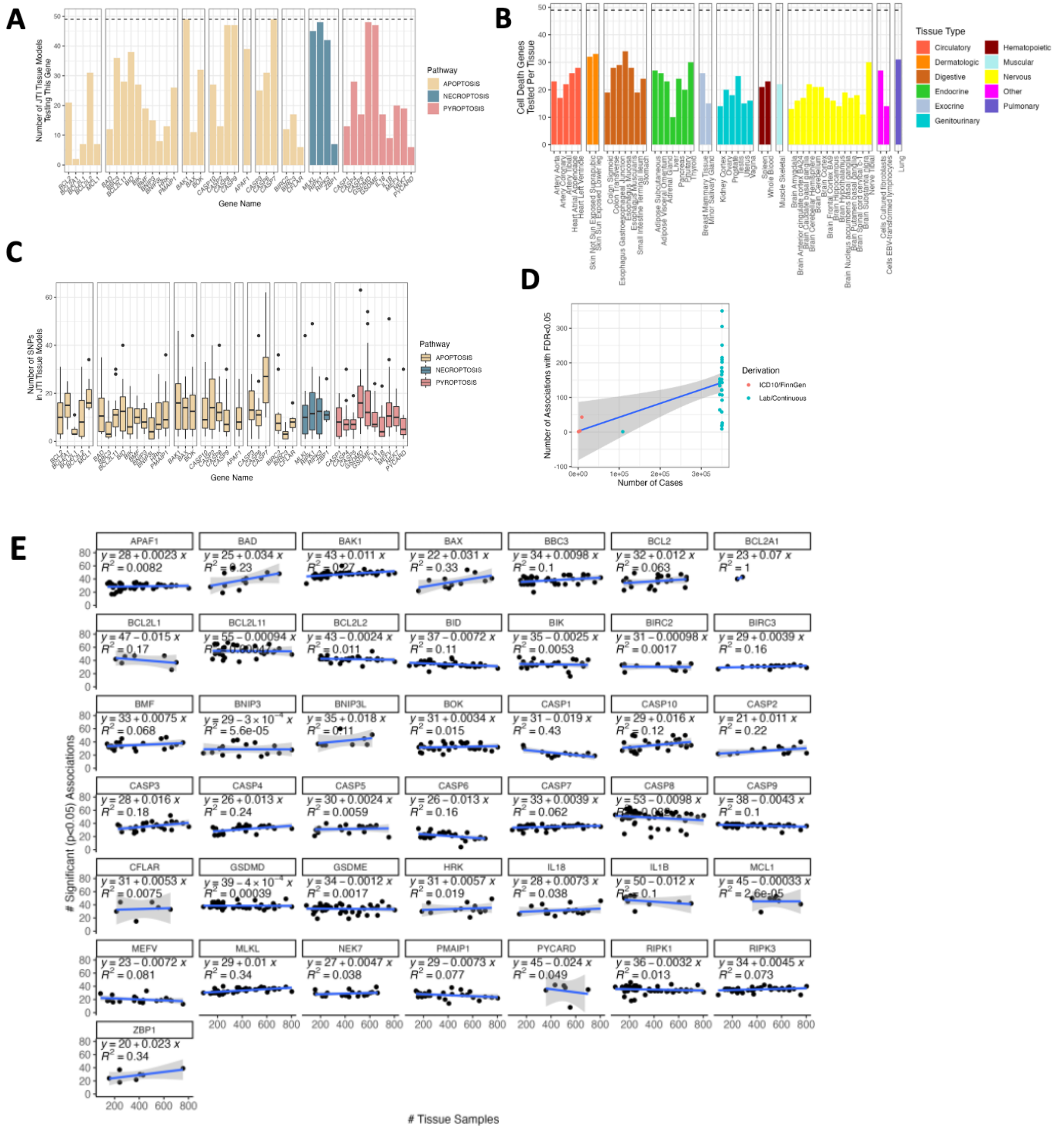


**Supplemental Figure 2: Joint Tissue Imputation Modeling and phenome wide scan results.**

2A: Number of tissues for which cell death genes were included in JTI association testing weights varies by gene.

2B: Number of cell death genes tested in each tissue.

2C: Distribution, across all tissues for which a given cell death gene is modeled, of the number of SNPs (eQTLs) included in JTI models.

2D: Relationship between the number of cases available for significant traits and the number of associations with FDR<0.05.

2E: For each gene in the cell death array, the correlation between the number of GTEx samples used to generate weights for each tissue and the number of significant ($p<0.05$) associations identified in that gene/tissue combination.
